# A Prospective Cohort Study Towards Improving Enhanced Recovery After Cesarean (ERAC) Pathways

**DOI:** 10.1101/2021.11.07.21265950

**Authors:** Liviu Cojocaru, Autusa Pahlavan, Suzanne Alton, Martha Coghlan, Hyunuk Seung, Ariel Trilling, Bhavani S Kodali, Sarah Crimmins, Katherine Goetzinger

## Abstract

**Objective:** To evaluate whether Enhanced Recovery After Cesarean (ERAC) pathways reduces inpatient and outpatient opioid use, pain scores and improves the indicators of postoperative recovery.

**Study design:** This is a prospective cohort study of all patients older than 18 undergoing an uncomplicated cesarean delivery (CD) at an academic medical center. We excluded complicated CD, patients with chronic pain disorders, chronic opioid use, acute postpartum depression, or mothers whose neonate demised before their discharge. Lastly, we excluded non-English and non-Spanish speaking patients. Our study compared the outcomes in patients before (pre-ERAC) and after (post-ERAC) implementation of an ERAC pathways. Primary outcomes were inpatient morphine milligram equivalent (MME) use and the patient’s delta pain scores. Secondary outcomes were outpatient MME prescriptions as well as indicators of postoperative recovery.

**Results:** Of 308 patients undergoing CD from October 2019 to September 2020, 196 were enrolled in the pre-ERAC cohort and 112 in the post-ERAC cohort. Patients in the post-ERAC cohort were less likely to require opioids in the postoperative period compared to the pre-ERAC cohort (35.7% vs. 18.4%, p<0.001). In addition, there was a significant reduction in the MME per stay in this cohort [16.8 MME (11.2-33.9) vs. 30 MME (20-49), p<0.001]. In the post-ERAC cohort, there was also a reduction in the number of patients who required prescribed opioids at the time of discharge (86.6 vs. 98%, p<0.001) as well as in the amount of MMEs prescribed [150 MME (112-150) vs. 150 MME (150-225), p<0.001; different shape of distribution]. Patients in the post-ERAC cohort had lower delta pain scores [2.2 (1.3-3.7) vs. 3.3 (2.3-4.7), p<0.001].

**Conclusion:** Our study has illustrated that our ERAC pathways reduced inpatient and outpatient opioid use as well as patient-reported pain scores while improving indicators of postoperative recovery.

## Introduction

Cesarean delivery (CD) is one of the most common surgical procedures in the United States^1^. Given patient expectations and misconceptions regarding the safety of labor, the medico-legal environment, and the limitations of modern technologies, the rate of CD is unlikely to decrease in the near future^2,3^. Consequently, standardizing care remains a top priority in health care quality and safety initiatives^4^. Enhanced Recovery After Surgery (ERAS) or, more specifically for obstetrics, Enhanced Recovery after Cesarean (ERAC) pathways have emerged as a multidisciplinary standardized bundled care approach to improve maternal outcomes^5^. Furthermore, there is evidence that ERAS help to address racial disparities by standardizing care^6^. Despite this, ERAS pathways have not been fully embraced in obstetrics^7^, leaving significant space for improvement^8^. Moreover, most of the studies have not extended ERAC pathways to specific populations such as opioid-naive patients, patients with postpartum depression, or patients receiving Magnesium Sulfate, allowing aforementioned confounders to affect the magnitude of the measured outcome.

Persistent opioid use in opioid-naive women after cesarean has become an alarming trend, where 1:50 to 1:300 women are at risk of chronic opioid use ^9,10^. Substance abuse has been cited as a major risk factor for pregnancy-associated deaths ^11,12^. Additionally, rising opioid use in pregnancy consequentially results in an increased incidence of neonatal abstinence syndrome^13^. Multimodal analgesia has been shown to decrease opioid use after cesarean^14^. However, non-opioid medications such as Gabapentin have not been yet incorporated into standard practice, despite data suggesting reduced postoperative opioid use ^15,16^ and endorsement by American College by Obstetricians and Gynecologists^17^.

Beyond opioid use, engaging patients in early ambulation and feeding has an additional positive impact on recovery^18–20^. A distinct difference in the obstetric population that may interfere with early ambulation and feeding is the postpartum Magnesium Sulfate use in patients with preeclampsia.

We aimed to reduce inpatient opioid use, outpatient opioid use, and reduce pain scores while improving the indicators of postoperative recovery and maximizing patient education. Furthermore, the specifics of our protocol are intended to decrease the knowledge gaps in ERAC pathways.

## Methods

This is a prospective cohort study of all patients older than 18 undergoing an uncomplicated cesarean delivery (CD) at an academic medical center. The Institutional Review Board at university of Maryland, Baltimore, approved the study under the protocol HP-00088872. The article was prepared following Reporting on ERAS Compliance, Outcomes, and Elements Research (RECOvER)^21^ and Strengthening the Reporting of Observational Studies in Epidemiology (STROBE)^22^. Data were collected and entered into a secure database, Research Electronic Data Capture (REDCap) software^23,24^, hosted by our institution.

Our study compared the outcomes in patients before (pre-ERAC) and after (post-ERAC) implementation of the ERAC pathways. Implemented core measures are described below, while the full ERAC pathways are available in Appendix 1.

Based on the reported information from clinical informatics on average opioid use in patients after cesarean, power analysis revealed that in order to detect a 50 % decreased in MME use per stay (90% chance) at a p < 0.05 statistical significance level, 112 patients in each cohort would be necessary.

Patient enrollment pre-ERAC occurred from October 2019 until February 2020. During this time period, the ERAC pathways were created, and all the necessary logistics for the implementation were prepared. The interventions were designed in accordance with Enhanced Recovery After Surgery (ERAS) Society guidelines for CD^20,25,26^. In March of 2020, the ERAC pathways were implemented on our Obstetric Care Unit (OBCU). We allowed a “wash-out period” of two months to permit staff adjustment and implementation of this new protocol. Following this period, post-ERAC enrollment occurred from May 2020 until September 2020.

Patient satisfaction surveys were administered during both pre- and post-ERAC periods as part of the implementation.

Exclusion criteria included CD done under general anesthesia, those complicated by massive transfusion events (defined as transfusion of six or more PRBC’s), bowel injury, CD requiring recovery in the intensive care unit, and skin incision other than Pfannenstiel. It is thought that these factors may result in higher opioid use and protracted recovery. Additionally, to further reduce the factors that can result in increased opioid use, we excluded patients with chronic pain disorders, chronic opioid use, acute postpartum depression, or mothers whose neonate demised before their discharge. Since we routinely provided patient recovery questionnaires in English and Spanish, we excluded patients who did not speak one of the two languages.

We collected: patient’s demographic information; past obstetrical, medical, and surgical history; the presence of mental health disorders and substance abuse (including urine toxicology results which is sent routinely upon admission); delivery information, delivery complications (chorioamnionitis, uterine incision extension, blood transfusion) and surgical complications (surgical site infection, postpartum hemorrhage, ileus or small bowel obstruction); neonatal information; inpatient opioid use along with pain scores as well as the amount of prescribed opioids; date and time of the ERAC interventions as described below in the secondary outcomes; and lastly, follow up data on postpartum follow up and readmission.

### ERAC team

Our multidisciplinary team consisted of representatives from obstetrics, obstetric anesthesiology, nursing leaders, patient-controlled analgesia services, pharmacy, project development, food and hospitality services, clinical informatics, data and analytics, clinical practices and professional development, materials management, patient experience advocacy, and other ancillary services.

### Implementation

Team leaders met regularly to discuss the ERAC pathways’ interventions, logistics of the implementation, the progress of the project, as well as the staff and patient feedback. The ERAC leaders were also responsible for staff educations. Physician education was done primarily at the Obstetrics & Gynecology department meetings. Nursing education was completed at the monthly and quarterly nursing meetings and on an individual basis. Moreover, education continued during other events, ranging from morning and evening sign-outs to email communications and grand rounds.

### Electronic medical records

A substantial change was made in our electronic medical record system (Epic Hyperspace®) to accommodate the implementation of our ERAC pathways into our clinical practice. All existing cesarean section order sets were reviewed and updated to include all new changes. An ERAC flowsheet was created to provide easy access to the MME usage and pain scores. This flowsheet also was used to evaluate the patient’s pain control and the quantity of outpatient opioid prescriptions. In addition, the flowsheet simplified nursing entry of all ERAC interventions.

### Core measure

Current practice was reviewed, and deficiencies were analyzed. Based on this review, a formal ERAC pathways were implemented. The focus was placed on essential and deficient areas. Four main areas were identified to require changes: multimodal analgesia, preoperative fasting time, early ambulation, and patient education. Details of each pathway are available in Appendix 1.

1. Routine use of Gabapentin was added to the multimodal analgesia protocol. The addition took place after a review of the available literature and consultation with neonatology and lactation services to confirm safety profiles.
2. Fasting before CD was reduced to six hours for light meals (examples are given in both patient’s education guide and instructional video) and two hours for fluids. A carbohydrate drink (50 grams) was routinely administered two hours before CD in non-diabetic mothers. After CD, the default diet was clear fluids and crackers within one hour and a light meal within four hours of arrival to the Post Anesthesia Care Unit (PACU).
3. The goals for ambulation were standardized to include specific instruction rather than “early ambulation was encouraged”. The goals were set for 0-24 hours(h), 24-48h, and more than 48h. The goals were adjusted based on clinical circumstances (i.e., patients requiring Magnesium Sulfate).
4. A paramount component of our ERAC pathways was patient education. We wanted to improve not only patient recovery but also their autonomy. The details of our ERAC program and goals of care were included in a patient education guide and instructional video that are available in English and Spanish at http://umm.edu/ERAC. The reading grade level was fourth grade for the patient video and fifth grade for the patient education guide (assessed with Flesch-Kincaid grading). This was in accordance with the Health Literacy guidelines for patient education materials. The patient education guide was provided during prenatal care visits as well as preoperative assessment. Those who missed education during prenatal care were educated on the obstetrical care unit during their preoperative visit. Video distribution was available at the outpatient clinical sites and all hospital patient rooms through our Telephone Initiated Guided Response video system.

### Compliance

Compliance was established at three levels:

1. ERAC champions were identified and were responsible for providing staff support and supervising compliance in the OBCU.
2. With the creation of the ERAC flowsheet, electronic data extraction of MME use and pain scores as well as secondary outcomes were periodically performed by the clinical informatics team at the request of the ERAC leaders. This information was used to assess compliance with documentation and assess the ERAC pathways’ efficacy.
3. Random charts were reviewed in detail to assess compliance with documentation.

### Outcomes

The primary outcomes were inpatient morphine milligram equivalent (MME) use and patient-reported pain scores. The MME was calculated per hospital day and per entire hospital stay. Pain was reported as delta pain (ΔP) and represented the difference between the patient’s reported pain and the patient’s pain goal.

Secondary outcomes were outpatient MME prescription, fasting time for liquids and solids (fasting time before surgery), time to feeding (the time to first feed after arrival to PACU), time to indwelling urinary catheter removal, time to ambulation, and time to hospital discharge.

### Statistical analysis

Descriptive statistics were used for frequency, median, and mean. Baseline demographics and outcomes were compared between pre-ERAC and post-ERAC cohorts using χ2 test or Fisher’s exact test for categorical variables and the Wilcoxon rank-sum test or Student’s t-test for continuous variables. Normality of distribution was established using the Kolmogorov-Smirnoff test. The associations between ERAC implementation and inpatient MME use and delta pain score for each postoperative day were measured using the Wilcoxon rank-sum test. Analysis was then stratified by Magnesium Sulfate use, maternal obesity, depression, and neonatal disposition (newborn nursey vs. neonatal intensive care unit), as these parameters may affect post-surgical goals. A negative binomial mixed model was adopted to solve right-skewed and over-dispersion of inpatient MME use. This model was used to evaluate the associations between inpatient MME use and ERAC implementation as well as to adjust for covariates. The goodness-of-fit Pearson χ2 was evaluated for the model fit of the negative binomial mixed model. The least square means were computed for postoperative days 0 to 4 to detect any differences in the inpatient MME use. A multinomial logistic regression model was conducted to control for potential confounders, identified either historically or through bivariate analysis. The odds ratio with a 95% confidence interval (CI) was used to measure the magnitude of the association. The variance inflation factor (VIF) and correlation coefficients were used to identify multicollinearity. The Hosmer-Lemeshow test and the area under the receiver operating characteristics (ROC) curve were used to assure goodness-of-fit and the discriminatory power for the logistic model. Analyses were performed with SAS software version 9.4 (SAS Institute, Cary, NC).

## Results

A total of 308 patients undergoing CD were enrolled in the study, 196 in the pre-ERAC cohort and 112 in the post-ERAC cohort. Baseline characteristics were comparable with regards to age, race and ethnicity, gravity, parity, and most pre-existing medical conditions. There was a higher incidence of diabetes mellitus in the pre-ERAC cohort (12.8 vs. 3.6%, p=0.008). The rate of obesity was also higher in the post-ERAC cohort [35.8 (30.9-41) vs. 33.3 (28.3-40.2), p<0.001] (Table 1). Similarly, the rate of prior laparotomy history other than CD was higher in the post-ERAC cohort (9.8% vs. 4.1%, p=0.04). Indication for CD was similar between cohorts, except that the post-ERAC cohort had a lower incidence of delivery for placental abnormality (0.9% vs. 6.1%, p=0.04). There was no difference in CD time, quantitative blood loss, or delivery and surgical complications between the two cohorts (Table 2 and 3). In addition, there was no difference in neonatal birth weight, Apgar score, or umbilical cord blood gases (Supplemental Table 1).

**Table 1.** Demographic and medical characteristics.

|  | **Total**<br>**(n=308)** | **preERAC**<br>**(n=196)** | **postERAC**<br>**(n= 112)** | **p-value** |
| --- | --- | --- | --- | --- |
| **Age, mean(SD)** | 30.6(6.1) | 30.6(6.4) | 30.7(5.5) | 0.9 |
| **Race, n(%)** |  |  |  | 0.2 |
| Hispanic | 17(5.5) | 7(3.6) | 10(8.9) |  |
| non-Hispanic Black | 195(63.3) | 121(61.7) | 74(66.1) |  |
| non-Hispanic White | 81(26.3) | 56(28.6) | 25(22.3) |  |
| Asian | 10(3.3) | 8(4.1) | 2(1.8) |  |
| other | 5(1.6) | 4(2.0) | 1(0.9) |  |
| **BMI, median(IQR)** | 33.3(28.3, 40.2) | 32.1(27.5, 39.5) | 35.8(30.9, 41) | 0.0002 |
| **Gravida, median(IQR)** | 3(2, 4) | 3(2, 4) | 3(2, 4) | 0.3 |
| **Para, median(IQR)** | 1(0, 2) | 1(0, 2) | 1(0, 1.5) | 0.1 |
| **Aborta, median(IQR)** | 1(0, 2) | 1(0, 2) | 0.5(0, 1) | 0.6 |
| **Medical problems** |  |  |  |  |
| **DM, n(%)** | 29(9.4) | 25(12.8) | 4(3.6) | 0.008 |
| **HTN, n(%)** | 61(19.8) | 36(18.4) | 25(22.3) | 0.4 |
| **Asthma, n(%)** | 67(21.8) | 42(21.4) | 25(22.3) | 0.9 |
| **CKD, n(%)** | 2(0.7) | 2(1.0) | 0 | 0.5 |
| **PIH, n(%)** | 61(19.8) | 45(23.0) | 16(14.3) | 0.07 |
| **GDM, n(%)** | 20(6.5) | 7(3.6) | 13(11.6) | 0.006 |
| **Cardiovascular disease, n(%)** | 7(2.3) | 4(2.0) | 3(2.7) | 0.7 |
| **Autoimmune, n(%)** | 4(1.3) | 2(1.0) | 2(1.8) | 0.6 |
| **Neurologic disease, n(%)** | 12(3.9) | 6(3.1) | 6(5.4) | 0.4 |
| **Thyroid disorder, n(%)** | 5(1.6) | 3(1.5) | 2(1.8) | 1.0 |
| **MDD, n(%)** | 69(22.5) | 48(24.5) | 21(18.9) | 0.3 |
| on medications, n(%) | 22(31.9) | 17(35.4) | 5(23.8) | 0.3 |
| positive toxicology, n(%) | 31(10.1) | 18(9.2) | 13(11.6) | 0.5 |
| **Other medical conditions, n(%)** | 17(5.5) | 9(4.6) | 8(7.1) | 0.3 |

ERAC: enhanced recovery after cesarean; BMI: body mass index; DM: diabetes mellitus; HTN: hypertension; GDM: gestational diabetes mellitus; MDD: major depressive disorder; SD: standard deviation; IQR: interquartile range; n: number.

**Table 2.** Surgical characteristics.

|  |  | **Past surgical history** |  |  | ***p*-value** |
| --- | --- | --- | --- | --- | --- |
|  |  | **Total**<br>**(n=308)** | **preERAC**<br>**(n=196)** | **postERAC**<br>**(n= 112)** |  |
| **No prior surgery, n(%)** |  |  |  |  | 0.5 |
|  | no | 154(50) | 95(48.5) | 59(52.7) |  |
|  | yes | 154(50) | 101(51.5) | 53(47.3) |  |
| **CS, n(%)** |  | 144(46.8) | 91(46.4) | 53(47.3) | 0.9 |
|  | number of CS, n(%) |  |  |  | 0.4 |
|  | 1 | 96(66.7) | 59(64.8) | 37(69.8) |  |
|  | 2 | 33(22.9) | 24(26.4) | 9(17.0) |  |
|  | 3 | 14(9.7) | 7(7.7) | 7(13.2) |  |
|  | 4 | 1(0.7) | 1(1.1) | 0 |  |
| **Laparotomy, n(%)** |  | 19(6.2) | 8(4.1) | 11(9.8) | 0.04* |
|  | number of laparotomies, n(%) |  |  |  | 1.0 |
|  | 1 | 16(84.2) | 8(100) | 8(72.7) |  |
|  | 2 | 1(5.3) | 0 | 1(9.1) |  |
|  | 3 | 1(5.3) | 0 | 1(9.1) |  |
|  | 4 | 1(5.3) | 0 | 1(9.1) |  |
| **Indication for CS** |  |  |  |  |  |
| **Elective, n(%)** |  | 102(33.1) | 59(30.1) | 43(38.4) | 0.1 |
| **Arrest of labor, n(%)** |  | 40(13.0) | 22(11.2) | 18(16.1) | 0.2 |
| **NRFHT, n(%)** |  | 64(20.8) | 39(19.9) | 25(22.3) | 0.6 |
| **Malpresentation, n(%)** |  | 35(11.4) | 24(12.2) | 11(9.8) | 0.5 |
| **Maternal indication, n(%)** |  | 53(17.2) | 37(18.9) | 16(14.3) | 0.3 |
| **Placental abnormality, n(%)** |  | 13(4.2) | 12(6.1) | 1(0.9) | 0.04* |
| **Other indication, n(%)** |  | 12(3.9) | 8(4.1) | 4(3.6) | 1.0 |
| **Blood loss, median(IQR)** |  | 759(520, 1067) | 763(507, 1047) | 751(522, 1123) | 0.9 |

ERAC: enhanced recovery after cesarean; CS: cesarean section; NRFHT: non-reassuring fetal heart tracing; IQR: interquartile range; n: number.

**Table 3.** Delivery and surgical complications.

| **Delivery complications** |  |  |  |  |
| --- | --- | --- | --- | --- |
|  | **Total**<br>**(n=308)** | **preERAC**<br>**(n=196)** | **postERAC**<br>**(n= 112)** | ***p*-value** |
| **No delivery complication, n(%)** |  |  |  | 0.2 |
| no | 46(14.9) | 25(12.8) | 21(18.8) |  |
| yes | 262(85.1) | 171(87.2) | 91(81.3) |  |
| **Chorioamnionitis, n(%)** | 14(4.6) | 9(4.6) | 5(4.5) | 1.0 |
| **Extension of the uterine incision, n(%)** | 13(4.2) | 11(5.6) | 2(1.8) | 0.1 |
| **Transfusion of blood products, n(%)** | 9(2.9) | 5(2.6) | 4(3.6) | 0.7 |
| **Other complications, n(%)** | 19(6.2) | 5(2.6) | 14(12.5) | 0.0005* |
| **Surgical complications** |  |  |  |  |
| **No surgical complications, n(%)** |  |  |  | 0.7 |
| no | 101(32.8) | 63(32.1) | 38(33.9) |  |
| yes | 207(67.2) | 133(67.9) | 74(66.1) |  |
| **SSI, n(%)** | 4(1.3) | 3(1.5) | 1(0.9) | 1.0 |
| **PPH, n(%)** | 97(31.5) | 62(31.6) | 35(31.3) | 0.9 |
| **SBO, n(%)** | 0 | 0 | 0 | 1.0 |
| **Other surgical complication, n(%)** | 11(3.6) | 8(4.1) | 3(2.7) | 0.8 |

ERAC: enhanced recovery after cesarean; SSI: surgical site infection; PPH: postpartum hemorrhage; SBO: small bowel obstruction; IQR: interquartile range; n: number.

### Morphine Milligram Equivalents used

There was a higher percentage of no postpartum opioids use in the post-ERAC cohort compared to the pre-ERAC cohort (35.7% vs. 18.4%, p<0.001). Furthermore, there was a significant reduction in the MME per day (postoperative days 1 to 4) and per stay [16.8(11.2-33.9) vs. 30(20-49), p<0.001] in the post-ERAC cohort (Table 4.1). The magnitude of difference was reduced in patients with depression and was only statistically significant for postoperative days 1 to 2. Consequentially, in patients with depression, there was a smaller difference in MME use per stay as well [17.8(8.4-38) vs. 36.3(25.2-54.8), p=0.03] (Table 4.1).

**Table 4.1.** Distribution of no postpartum MME use.

|  | MME, median(IQR) |  |  |  |  |  |
| --- | --- | --- | --- | --- | --- | --- |
|  | Day 0<br>(n=308) | Day 1<br>(n=307) | Day 2<br>(n=307) | Day 3<br>(n=230) | Day 4<br>(n=61) | Overall average<br>(n=308) |
| **Total** | 0(0, 0.6) | 15(0,30) | 15(0, 37.5) | 15(0, 30) | 15(0, 30) | 12.5(0.6, 28.4) |
| **No MME use, n(%)** | 229(74.4) | 102(33.2) | 105(34.2) | 82(35.7) | 17(27.9) | 76(24.7) |
| **preERAC** | (n=196) | (n=195) | (n=195) | (n=160) | (n=45) | (n=196) |
|  | 0(0, 6.8) | 22.5(5, 45) | 22.5(0, 52.5) | 15(0, 33.8) | 15(7.5, 30) | 20.5(5.6, 35.2) |
| **No MME use, n(%)** | 141(71.9) | 47(24.1) | 51(26.2) | 41(25.6) | 9(20) | 36(18.4) |
| **postERAC** | (n=112) | (n=112) | (n=112) | (n=70) | (n=16) | (n=112) |
|  | 0(0, 0) | 3.8(0, 15) | 7.5(0, 15) | 0(0, 15) | 3.8(0, 18.8) | 4.3(0, 15.4) |
| **No MME use, n(%)** | 88(78.6) | 55(49.1) | 54(48.2) | 41(58.6) | 8(50) | 40(35.7) |
| **p-value** | 0.2 | <0.0001* | <0.0001* | <0.0001* | 0.03* | <0.0001* |
|  | MME, median(IQR) in patients with MDD |  |  |  |  |  |
| Patient with MDD | Day 0<br>(n=69) | Day 1<br>(n=68) | Day 2<br>(n=68) | Day 3<br>(n=55) | Day 4<br>(n=14) | Overall average<br>(n=69) |
| **Total** | 0(0, 25.2) | 15(3.7, 45.5) | 22.5(5, 45) | 15(0, 30) | 15(7.5, 30) | 18(15, 39.4) |
| **No MME use, n(%)** | 44(63.8) | 16(23.5) | 16(23.5) | 17(30.9) | 3(21.4) | 11(15.9) |
| **preERAC** | (n=48) | (n=47) | (n=47) | (n=39) | (n=11) | (n=48) |
|  | 0(0, 26.3) | 30(15, 60) | 30(10, 52.5) | 15(7.5, 30) | 15(7.5, 30) | 25.3(7.9, 47.0) |
| **No MME use, n(%)** | 29(60.4) | 8(17.0) | 9(19.2) | 8(20.5) | 1(9.1) | 6(12.5) |
| **postERAC** | (n=21) | (n=21) | (n=21) | (n=16) | (n=3) | (n=21) |
|  | 0(0, 7.5) | 7.5(0, 15) | 7.5(0, 22.5) | 15(0, 11.3) | 0(0, 45) | 8.4(1.5, 17.5) |
| **No MME use, n(%)** | 15(71.4) | 8(38.1) | 7(33.3) | 9(56.3) | 2(66.7) | 5(23.8) |
| **p-value** | 0.4 | 0.007* | 0.005* | 0.009* | 0.3 | 0.01* |

ERAC: enhanced recovery after cesarean; MDD: major depressive disorder; MME: morphine milligram equivalent; IQR: interquartile range; n: number.

**Table 4.2.** Distribution of pain score.

|  | Delta pain score, median(IQR) |  |  |  |  |  |
| --- | --- | --- | --- | --- | --- | --- |
|  | **Day 0**<br>**(n=299)** | **Day 1**<br>**(n=308)** | **Day 2**<br>**(n=307)** | **Day 3**<br>**(n=264)** | **Day 4**<br>**(n=78)** | **Overall average**<br>**(n=308)** |
| **Total** | 1.8(0.6, 3.2) | 3.3(1.9, 5) | 3.8(2, 5) | 3.3(1.6, 5) | 3.3(2, 5.4) | 3.0(1.7, 4.4) |
| **preERAC** | **(n=196)** | **(n=196)** | **(n=195)** | **(n=182)** | **(n=54)** | **(n=196)** |
|  | 1.7(0.6, 3.3) | 3.8(2.6, 5.1) | 4.1(2.4, 5.3) | 3.9(2.3, 5.5) | 3.6(2.7, 5.9) | 3.3(2.3, 4.7) |
| **postERAC** | **(n=103)** | **(n=112)** | **(n=112)** | **(n=82)** | **(n=24)** | **(n=112)** |
|  | 2(0.8, 3) | 2(1, 4) | 3(1, 4.3) | 2(1, 4) | 2.3(0.5, 4) | 2.2(1.3, 3.7) |
| **p-value** | 0.7 | <0.0001* | 0.0001* | <0.0001* | 0.005* | <0.0001* |

ERAC: enhanced recovery after cesarean; MDD: major depressive disorder; MME: morphine milligram equivalent; IQR: interquartile range; n: number.

The number of patients who required prescribed opioids at the time of discharge was significantly decreased in the post-ERAC cohort (86.6 vs. 98%, p<0.001). The post-ERAC patients also had fewer MMEs prescribed [150 (112-150) vs. 150 (150-225), p<0.001; different shape of distribution] compared to the pre-ERAC cohort (Table 5).

**Table 5.**
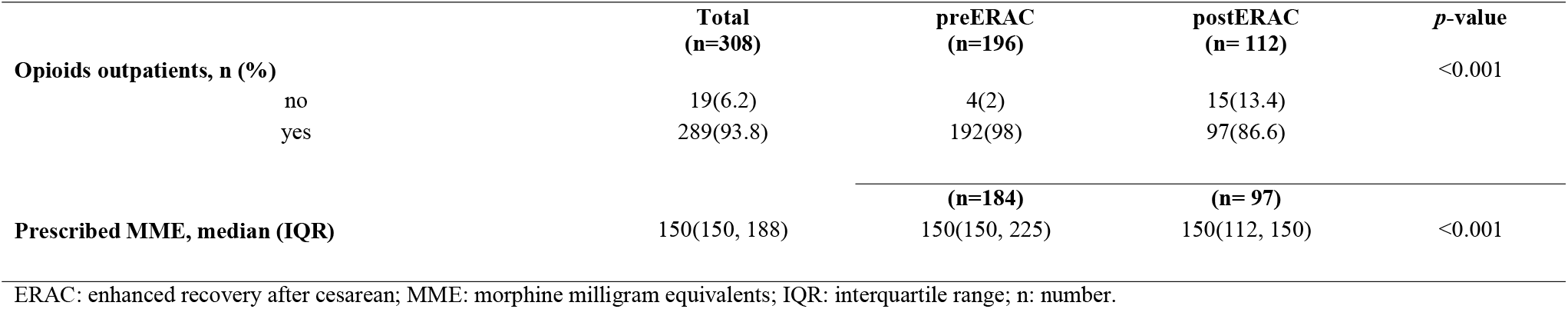
Distribution of prescribed outpatient opioids.

### Delta pain scores

The patients in the post-ERAC cohort had lower delta pain scores on postoperative days 1 to 4 as well as lower overall delta pain scores [2.2 (1.3-3.7 vs. 3.3 (2.3-4.7), p<0.001] (Table 4.2).

### Secondary outcomes (Table 6.1)

**Table 6.1.**
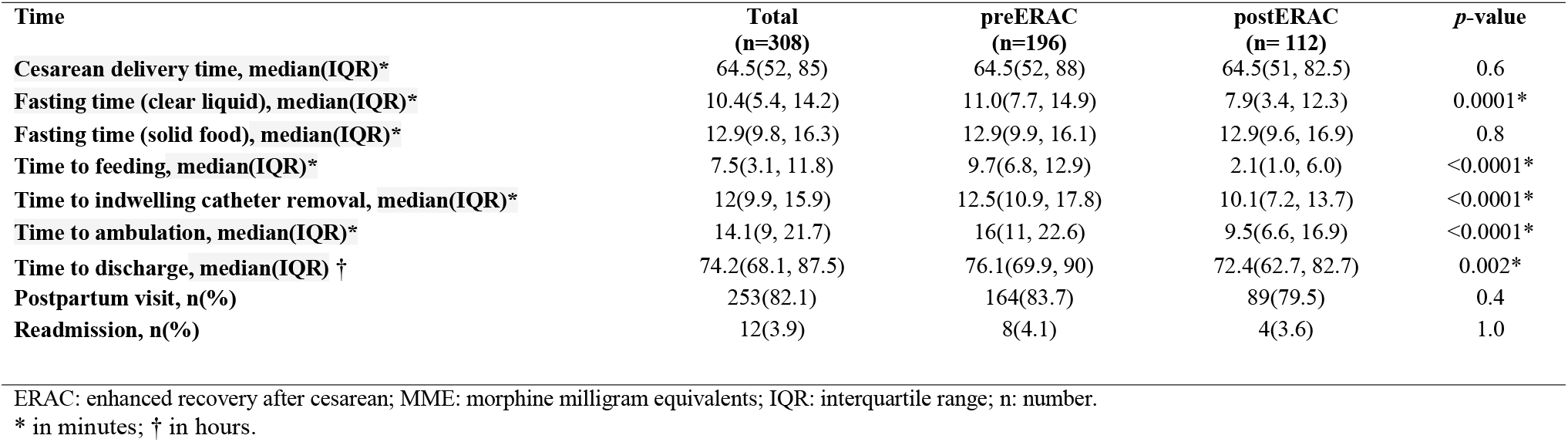
Recovery outcomes and follow up.

**Table 6.2.** Time to ambulation and discharge adjusted for Magnesium Sulfate use neonatal NICU admission.

| **Time** | **Magnesium Sulfate** |  |  | **No Magnesium Sulfate** |  |  |
| --- | --- | --- | --- | --- | --- | --- |
|  | **preERAC**<br>**(n=32)** | **postERAC**<br>**(n=14)** | ***p*-value** | **preERAC**<br>**(n=161)** | **postERAC**<br>**(n=98)** | ***p*-value** |
| **Time to ambulation,**<br>**median(IQR)** | 25.7(24.5, 26.6) | 25.4(22.7, 28.1) | 0.6 | 14.4(10.5, 19.1) | 9.3(6.5, 14.5) | <0.0001* |
| **Time to discharge,**<br>**median(IQR)** | 86.1(73.9, 96.2) | 76.4(69.8, 108.9) | 0.7 | 74.9(69, 83.9) | 72.1(61.7, 79.7) | 0.003* |
|  | **NICU admission** |  |  | **No NICU admission** |  |  |
|  | **preERAC**<br>**(n=76)** | **postERAC**<br>**(n=37)** | ***p*-value** | **preERAC**<br>**(n=112)** | **postERAC**<br>**(n=75)** | ***p*-value** |
| **Time to ambulation,**<br>**median(IQR)** | 17.1(10.5, 25.8) | 9.6(7.7, 19.2) | 0.006* | 15.4(11.1, 20.7) | 9.5(6.5, 16.4) | <0.0001* |
| **Time to discharge,**<br>**median(IQR)** | 78.7(69.3, 95.2) | 86(73.8, 96.4) | 0.2 | 75.2(70.1, 81) | 69(54, 74.3) | <0.0001* |

ERAC: enhanced recovery after cesarean; NICU: neonatal intensive care unit; IQR: interquartile range; n: number.

#### Fasting time

Preoperative fasting time for clear liquids was significantly less in the post-ERAC [7.9h (3.4, 12.3) vs. 11.0h (7.7, 14.9), p<0.001]. There was no significant difference in the fasting time for solids [12.9h (9.6, 16.9) vs. 12.9h (9.9, 16.1), p=0.8]. However, time to feeding postoperatively occured significantly sooner in the post-ERAC cohort [2.1h (1.0, 6.0) vs. 9.7h (6.8, 12.9), p<0.001].

#### Time to indwelling urinary catheter removal

The time to indwelling urinary catheter removal was statistically significant less in the post-ERAC cohort [10.1h (7.2,13.7) vs. 12.5h (10.9,17.8), p<0.001].

#### Time to ambulation

The time to ambulation was also statistically significant less in the post-ERAC cohort [9.5h (6.6,16.9) vs. 16.0h (11-22.6), p<0.001]. When we stratified by Magnesium Sulfate use, this difference persisted in those who did not receive Magnesium Sulfate [9.5h (6.6,16.9) vs. 16.0h (11,22.6), p<0.001]. In those who did receive post-operative Magnesium Sulfate, time to ambulation was similar between the two cohorts [25.4h (22.7,28.1) vs. 25.7h (24.5,26.6, p=0.6] (Table 6.2)..

#### Time to discharge

The time to discharge was statistically significant less in the post-ERAC cohort [72.4h (62.7, 82.7) vs. 76.1h (69.9,90), p=0.002]. When stratified by NICU admission and magnesium Sulfate use, the difference was not statistically significant (Table 6.2).

### Multivariate analysis

Inpatient MME used was noted to be right-skewed and over-dispersed. In order to achieve patterns in the data which would be more interpretable and meet the assumptions of inferential statistics, a negative binomial mixed model was conducted. Only statistically significant variables from the bivariate analysis were included (Supplementary Table 2). To help assess the fit of the model, we used the goodness-of-fit Pearson χ2. The χ2/DF was 0.48, which yielded a *p*-value of 1. The non-significant *p*-value suggests that the negative binomial model is a good fit for the data. The measure of over-dispersion dropped from 12.9 (Poisson model) to 0.48 (negative binomial model), indicating that over-dispersion was no longer a problem. After adjusting for confounders, patients in the post-ERAC cohort were still less likely to use opioids on postoperative days 1, 2, 3, and per total stay. Delivery complications were an independent predictor for increased opioid use (Supplementary Table 3).

Additionally, for each hour decrease in time to feeding, there was a 5% decrease in opioid use, while for each unit increase in pain scores, patients were twice as likely to use opioids. For outpatient opioid use, we categorized prescribed MME based on a 150 cut-off due to frequency of distribution. We adjusted for BMI, race/ethnicity, depression, toxicology results, inpatient MME use, time to discharge, and pain goals. The overall model was significant (likelihood χ2 (11) = 22.9 with a p=0.02) and pseudo *R*^2^ of 12.7%. The Hosmer and Lemeshow χ2 (8) statistics was 9.1, with a *p*-value of 0.3. The area under the receiver-operating curve (AUC) was 0.68, denoting a fair model fit. Being in the post-ERAC cohort was the only predictor of decreased inpatient MME use [OR 0.38 (0.18-0.76), p=0.006]. The patients in the post-ERAC cohort were 38% less likely to be prescribed nore than 150 MME compared to pre-ERAC cohort.

## Discussion

### Principal findings

Implementation of our ERAC pathways have significantly reduced inpatient and outpatient opioid use while reducing patient pain scores. Although the relative reduction in opioid use might appear lower than in other studies, our study demonstrated either a more substantial absolute reduction in opioid use was more substantial^27^ or a greater reduction in the pain scores ^14^. Furthermore, we were able to implement Gabapentin as part of our standard pain management algorithm with no reported adverse events. Thus, we continue to use it as a part of our multimodal analgesia.

Preoperative fasting time for clear liquids was significantly reduced in our post-ERAC cohort, and although while the fasting time for solids remained unchanged pre and post-ERAC, this is likely related to the delays in the start of elective cases or policies related to feeding while laboring (e.g., clear liquids only if the patient is on Magnesium Sulfate). The benefits of oral intake in labor are related to physiological and psychological advantages^28^. Prolonged fasting with resulting ketosis and ketoacidosis has been found to result in prolonged labor, increased need for induction and augmentation in nulliparous women, forceps deliveries, and increased blood loss^29,30^. On the contrary, oral hydration in labor has been shown to decrease maternal urine ketones at delivery and may have a benefit on the uterus and placental perfusion^31^. To remove barriers for early ambulation, we planned early discontinuation of intravenous fluids and early removal of indwelling urinary catheters. In addition, since effective pain relief per se does not automatically lead to increased ambulation, we set up mobilization goals at 0 to 8 h, 8 to 24 h, and more than 24 h. Instructions are described in detail in Appendix 1. Although provider discretion was allowed for patients who received postpartum Magnesium Sulfate, feeding, early indwelling urinary catheter removal, and early mobilization were encouraged. This resulted in a significant reduction of time to feeding, time to removal of the indwelling urinary catheters, and time to ambulation. It has been demonstrated that early feeding results in improved maternal satisfaction, accelerated return of bowel function, reduced time to ambulation, and decreased length of stay^20^. Early removal of indwelling urinary catheters has been associated with a lower incidence of urinary tract infection, early ambulation, and decreased length of stay^19^. Consequently, early ambulation has the potential benefits of rapid return of bowel function, reduced risk of thromboembolism, and decreased length of stay^20^.

Finally, time to discharge was significantly reduced in the post-ERAC group; however, the clinical significance of this reduction is less clear. Factors other than maternal recovery influence the time of discharge, including neonatal monitoring and disposition. Nevertheless, several-hour differences in the time to discharge can affect insurance coverage beyond 72 hours, thereby affecting overall hospital cost^32^.

Most importantly, we made patient education the foundation of our protocol implementation. Since most patient education materials are written at reading levels considerably higher than that of the average United States adult^33^, we made it a priority to achieve an optimal reading level for our ERAC education materials(at or below 5^th^ grade)^34^. Patient education has become a cornerstone of successful ERAC implementation^8^, which starts during prenatal care, continues throughout hospitalization, and follows through the postpartum care. This is aimed to attend patients’ emotional needs that are often unrecognized in their struggle to regain control of their health and well-being^35^.

### Strengths

One of the main strengths is the prospective nature of our study. In addition, our pathways contain several elements that have not been previously reported:

1. Inclusion of non-elective CD. Most of the reported studies included elective CD, and only some urgent CD^36,37^, leaving out unscheduled non-urgent cases. Although multimodal pain regimens have been studied in the settings of non-elective CD^38^, they have not been applied along with ERAC pathways. Our algorithm can be applied across all uncomplicated CD categories, including those CDs performed during labor.
2. Integration of Gabapentin as part of the multimodal analgesia within the ERAC pathways. Gabapentin has been studied as an analgetic after CD, but not in the context of ERAC pathways^15,16^.
3. Use of a realistic approach to the patient pain goals (by studying the difference between the patient’s pain goals and the patient’s pain scores). While a previous study reported delta pain, that study regimen did not demonstrate an actual reduction in pain scores^27^.
4. Patient-centered implementation. A patient’s education guide and instructional video were created at the 4^th^ and 5^th^ reading level with approval from the patient’s advocacy department. The translation was done in Spanish to ensure inclusion of the Hispanic population. This population has been reported to have decreased pain control after CD^39^. Furthermore, we routinely offered patient satisfaction surveys to every patient on postoperative day one and the day of discharge.

### Limitations

One of the limitations of our study is the enrollment of only uncomplicated CD. Based on our results, it seems reasonable that this algorithm can be applied to complicated CD; however, our study was not designed to evaluate this population specifically. As with any quality improvement (QI) initiative, there is a potential for performance bias (after implementing the QI, the post-intervention cohort receives more attention during medical care). Another limitation is that we did not verify the opioid prescription dispensed from the pharmacy. Our study did not include long-term postoperative follow-up; therefore, we were unable to identify additional difficulties in recovery after discharge from the hospital. However, as part of our Obstetrical Transitional Care Program, patients receive phone calls from a dedicated nursing team assessing their recovery from CD on a routine basis. We did not identify more difficulties with pain control in the post-implementation period, nor did we note any difference in the re-admission rates. Finally, the study did not evaluate the cost analysis of our interventions, which will be a crucial next step in determining the success of these pathways in health care utilization.

## Conclusion

In conclusion, our study has illustrated that our ERAC pathways reduced inpatient and outpatient opioid use while reducing pain scores and improving indicators of postoperative recovery.

Specifically, we focused on patient education as being the foundation of our successful protocol implementation. Future efforts to expand these pathways to additional populations will be paramount as we move forward to streamline health care utilization while improving overall patient experience.

## Supporting information

STROBE checklist

RECOvER checklist

## Data Availability

All data produced in the present study are available upon reasonable request to the authors

## Acknowledgments

The authors are grateful to Dr. Christopher Harman for his unconditional support in implementing this project. We are thankful to Ms. Elizabeth Groncki, who greatly facilitated the project development. Also, we are appreciative of Tracy Keith’s and Cara Sullivan’s nursing leadership during the implementation of this project. Gena Stanek helped prepare the patient’s educational materials to the recommended reading level, and Alexandra Bessent translated it into an engaging patient’s guide and instructional video.

## Contribution to authorship

LC and KG drafted the manuscript. HS performed the statistical analysis and ensured its correct interpretation. All authors contributed to the writing, editing, and approving the final version.

## Tables

**Supplemental Table 1.** Neonatal Outcomes.

|  | **Total**<br>**(n=308)** | **preERAC**<br>**(n=196)** | **postERAC**<br>**(n= 112)** | ***p*-value** |
| --- | --- | --- | --- | --- |
| **Gestational age, median(IQR)** | 268(253, 275) | 268(251, 276) | 268(256, 274) | 0.7 |
| **Neonatal gender, n(%)** |  |  |  | 0.9 |
| male | 151(49.0) | 97(49.5) | 54(48.2) |  |
| female | 157(51.0) | 99(50.5) | 58(51.8) |  |
| **Birth weight, median(IQR)** | 3125(2510, 3470) | 3110(2370, 3455) | 3163(2667, 3525) | 0.1 |
| **Apgar at 1 min, median(IQR)** | 8(7, 8) | 8(7, 8) | 8(8, 8) | 0.8 |
| **Apgar at 5 mins, median(IQR)** | 9(9, 9) | 9(9, 9) | 9(9, 9) | 0.6 |
| **pH artery, median(IQR)** | 7.22(7.17, 7.26) | 7.22(7.17, 7.27) | 7.22(7.18, 7.26) | 0.4 |
| **BE artery, median(IQR)** | -4.15(-6.4, -2.9) | -4.15(-6.3, -2.75) | -4.2(-6.5, -2.9) | 0.8 |
| **pH vein, median(IQR)** | 7.29(7.24, 7.32) | 7.29(7.24, 7.33) | 7.29(7.25, 7.32) | 0.6 |
| **BE vein, median(IQR)** | -3.3(-5.1, -2.2) | -3.5(-5.3, -2.1) | -3.2(-4.7, -2.3) | 0.6 |

BE: base excess; IQR: interquartile range; n: number.

**Supplemental Table 2.** Bivariate analysis of the outcomes.

| Variables | Outcomes, median(IQR) |  |  |  |  |  |
| --- | --- | --- | --- | --- | --- | --- |
|  | Inpatient MME | Pain scores | Outpatient MME | Time to ambulation | Time to feeding | Time to discharge |
| **ERAC implementation** |  |  |  |  |  |  |
| preERAC | 20.5(5.6, 35.2) | 3.3(2.3, 4.7) | 150(150, 225) | 16(11, 22.6) | 9.7(6.8, 12.9) | 76.1(69.9, 90) |
| postERAC | 4.3(0, 15.4) | 2.2(1.3, 3.7) | 150(112.5, 150) | 9.5(6.6, 16.9) | 2.1(1, 6) | 72.4(62.7, 82.7) |
| **p-value** | <0.0001* | <0.0001* | <0.0001* | <0.0001* | <0.0001* | 0.002* |
| **Obesity** |  |  |  |  |  |  |
| non-obese | 14.4(2.4, 30) | 3.1(1.8, 4.4) | 150(150, 225) | 15.2(10.6, 19.6) | 9.1(4.7, 12.5) | 75.2(69, 87.1) |
| obese | 11.3(0, 28.2) | 3.0(1.7, 4.4) | 150(150, 150) | 13.3(8.7, 22) | 6.5(2.2, 11.8) | 73.8(68.1, 90.3) |
| **p-value** | 0.4 | 0.4 | 0.04* | 0.3 | 0.007* | 0.8 |
| **MDD** |  |  |  |  |  |  |
| no | 11.3(0, 26.3) | 2.9(1.7, 4.2) | 150(150, 187.5) | 14.1(9.1, 22) | 7.2(3.1, 11.7) | 74.3(67.8, 88.1) |
| yes | 18(5, 39.4) | 3.5(2.4, 5.0) | 150(150, 187.5) | 14.3(9, 20.2) | 8.7(2.6, 12) | 74.6(69.1, 88.7) |
| **p-value** | 0.02* | 0.005* | 0.9 | 0.9 | 0.4 | 0.6 |
| **Magnesium Sulfate use** |  |  |  |  |  |  |
| no | 11.6(1, 27) | 3(1.8, 4.4) | 150(150, 150) | 12.2(8.3, 17.6) | 7.5(3, 11.3) | 73.8(67.4, 83.4) |
| yes | 19.5(0, 33.8) | 2.8(1.6, 3.8) | 150(150, 225) | 25.7(24.4, 26.7) | 11.4(3.7, 14.8) | 83.9(71.9, 97.4) |
| **p-value** | 0.3 | 0.4 | 0.06 | <0.0001* | 0.03* | 0.0003* |

ERAC: enhanced recovery after cesarean; MME: morphine milligram equivalents; MDD: major depressive disorder; IQR: interquartile range; n: number.
\*statistically significant; p-values were computed using the Wilcoxon rank-sum test.

**Supplemental Table 3.** Multivariate analysis for inpatient MME.

| **Effect** | **Estimate (SE)** | **Rate ratio** | **95% confidence limit** | ***p*-value** |
| --- | --- | --- | --- | --- |
| postERAC vs. preERAC | -0.91(0.39) | 0.40 | (0.18, 0.87) | 0.02 |
| Day 1 vs. Day 0 | 1.14(0.22) | 3.13 | (2.04, 4.81) | <0.0001 |
| Day 2 vs. Day 0 | 1.25(0.23) | 3.49 | (2.22, 5.53) | <0.0001 |
| Day 3 vs. Day 0 | 0.84(0.24) | 2.32 | (1.44, 3.72) | 0.0005 |
| Delivery complications | 0.89(0.31) | 2.44 | (1.32, 4.46) | 0.004 |
| Time to feeding | -0.05(0.022) | 0.95 | (0.91, 0.99) | 0.002 |
| Overall average dela pain scores | 0.68(0.073) | 1.97 | (1.68, 2.28) | <0.0001 |

ERAC: enhanced recovery after cesarean; IQR: interquartile range; n: number.

