## Supplementary material for "A Prospective Cohort Study Towards Improving Enhanced Recovery After Cesarean (ERAC) Pathways": RECOvER checklist

**Supplementary Table 1. RECOvER Checklist.**

| | Item | Recommendation | Page |
|---|---|---|---|
| *Title* | | | |
| Title | 1 | A Prospective Cohort Study Towards Improving Enhanced Recovery After Cesarean (ERAC) Pathways. | 1 |
| *Introduction* | | | |
| Background | 2 | Use of Gabapentin as part of the multimodal regimen. Expansion of ERAC pathways to non-elective cesarean deliveries (CD). Evaluate the impact of Magnesium Sulfate use on the indicators of postoperative recovery. | 4 |
| Guidelines | 3 | Guidelines for antenatal, preoperative, intraoperative, and postoperative care in cesarean delivery. Enhanced Recovery After Surgery Society recommendations (Part 1,2, and 3). | References 20, 25, 26 |
| Outcomes | 4 | *Primary outcome*<br>Inpatient morphine milligram equivalents (MME) use and pain scores.<br>*Secondary outcomes*<br>Prescribed MME, fasting time, time to feeding, time to indwelling urinary catheter removal, time to ambulation, time to discharge. | 5 |
| *Methods* | | | |
| IRB approval | 5 | IRB HP-00088872. | 6 |
| Study design | 6 | Prospective cohort study following STROBE guidelines. | 6 |
| Setting | 7 | Single tertiary care academic institution with a stable group of surgeons during the study period. | 6 |
| Timing | 8 | Pre-implementation enrollment occurred from October 2019 until February 2020 and post-implementation from May 2020 until September 2020. Events were assessed daily from surgery to discharge. All patients followed until 6 weeks postpartum. | 7 |
| Participants | 9 | *Inclusion criteria*<br>Age older than 18, participating in ERAC pathways, undergoing CD.<br>*Exclusion criteria*<br>CD done under general anesthesia; CD complicated by massive transfusion events (defined as transfusion of six or more PRBC's), bowel injury, or requiring recovery in the intensive care unit; skin incision other than Pfannenstiel; patients with chronic pain disorders, chronic opioid use, acute postpartum depression; mothers whose neonate demised before their discharge; non-English or non-Spanish speaking patients. | 6-7 |

**Supplementary Table 1. RECOvER Checklist.**

| | Item | Recommendation | Page |
|---|---|---|---|
| Enhanced recovery protocol | 10 | ERAC pathways initiated in May 2020. | 7 |
| | 11 | Provide a flow diagram or table through the continuum of care detailing the enhanced recovery protocol, including the following elements: | Methods Appendix 1 |
| | | (a) Preadmission patient education regarding the protocol<br>Patient's education guide and instructional video were the cornerstones of the education. This was in addition to the routine education provided during the prenatal care, preoperative visits (for elective cases), and postoperative care, in accordance with the education and instructions provided to the medical care team. | 9 |
| | | (b) Preadmission screening and optimization as indicated for nutritional deficiency, frailty, anemia, HbA1c, tobacco cessation, and ethanol use.<br>The following were addressed during prenatal care. Ethanol levels are part of the hospital admission screening. Additionally, anemia, glucose control, tobacco smoking addressed during hospitalization. | 9 |
| | | (c) Fasting and carbohydrate loading guidelines<br>Light meal up to 6 hours before the CD. Clear liquids up to 2 hours before the CD. Carbohydrate loading with Ensure presurgical clear 2 hours before the CD. | 9 |
| | | (d) Preemptive analgesia (dose, route, timing)<br>*Preoperative*<br>Acetaminophen 1g per os (PO) and Gabapentin 600 mg PO at the time of presurgical drink.<br>*Intraoperative*<br>Ketorolac 15mg IV after peritoneum/ fascia is closed.<br>Local wound infiltration with 20 ml of 0.25% Ropivacaine.<br>Acetaminophen 1g per rectum if the patient has not received a preoperative dose. | 9 |
| | | (e) Anti-emetic prophylaxis (dose, route, timing)<br>*Preoperative*<br>Bicitra 30 ml preoperative. Famotidine 20mg IV pre-operatively.<br>Ondansetron 4 mg IV preoperative and Reglan 10mg IV pre-operatively.<br>If unable to administer Zofran or Reglan, substitute with Dexamethasone 4 mg IV after delivery of the neonate (in non-diabetic, immunocompetent mothers). | Appendix 1 |

**Supplementary Table 1. RECOvER Checklist.**

| Item | Recommendation | Page |
|---|---|---|
| | Transdermal scopolamine patch for patients at high risk of postoperative nausea or vomiting (delivers ~ 1mg over 72h).<br>I*ntraoperative*<br>Prophylactic vasopressor infusion (started at 10 mcg/min immediately after the intrathecal local anesthetic injection and titrated to blood pressure and pulse rate). | |
| | (f) Limited to less than 3 liters for routine cases. | N/A |
| | (g) Types, doses, and routes of anesthetics administered: low-medium dose long-acting opioid.<br>Intrathecal neuraxial anesthesia: Morphine 0.2 mg.<br>Epidural neuraxial anesthesia: Morphine 2 mg. | Appendix 1 |
| | (h) Patient warming strategy<br>Forced-air warming, intravenous fluid warming, increased operating room temperature (69 ˚F). | N/A |
| | (i) Management of postoperative fluids<br>Peripheral lock IV at 600 mL oral intake (for patients delivered after 8 pm, peripheral lock IV at 8 am the day after surgery if tolerating PO fluids).<br>For patients on PCA, intravenous fluids will be reduced to 10cc/hour once Oxytocin infusion is complete and the above criteria are met. | Appendix 1 |
| | (j) Postoperative analgesia and anti-emetic plans<br>Acetaminophen 1g PO every 6 hours. Ibuprofen 800mg PO every 8 hours. Gabapentin 300mg PO every 8 hours. Administration follows preoperative doses (intraoperative).<br>Reglan 10mg every 8 hours PRN for postoperative nausea and vomiting. | Appendix 1 |
| | (k) Plan for opioid minimization<br>*PRN opioids*<br>Tramadol 25-50mg PO every 6 hours PRN can be substituted for Acetaminophen or Ibuprofen (should the patient have contraindications to any of those).<br>Oxycodone 2.5mg PO for moderate pain and 5mg PO for severe pain every 4 hours PRN (once the patient is off PCA pump).<br>*PCA settings to reflect decreased dosing*<br>Hydromorphone loading: none. Demand dose: 0.2mg every 8 minutes (if patient weight <125kg) or every 10 minutes (if patient weight >125kg). Limit: 1.2mg per hour. | Appendix 1 |

**Supplementary Table 1. RECOvER Checklist.**

| | Item | Recommendation | Page |
|---|---|---|---|
| | | Goal to be discontinued at 8-12h postoperative. Patient needs to be tolerating PO. When transitioned to PO, the first dose will be administered by PCA services.<br>*Breakthrough pain*<br>During the first 18 hours following administration of regional long-acting opioid call anesthesia.<br>For patients on PCA with uncontrolled pain: 0.4mg every 10 minutes to a max of 5 doses per hour if done by a physician or two boluses per hour if done by an RN.<br>For patients not on PCA: Hydromorphone 0.5mg-2mg IV after evaluation by a physician. | |
| | | (l) Drain and line management<br>No routine wound drains. Removal of the indwelling urinary catheter at 6 hours postoperative or 2 hours after ambulation, whichever comes first. No later than 12h. For patients on Magnesium Sulfate, the indwelling urinary catheter will be maintained at the provider's discretion. | N/A |
| | | (m) Early mobilization strategy<br>*0-8h postoperative*<br>Sit on the edge of the bed at least once. Out of bed (OOB) to chair at least once. Ambulation as tolerated (room/ hallway). For patients on magnesium, mobilization will be limited to sitting on the edge of the bed/ OOB at least once.<br>*8-24h postoperative*<br>Ambulation as tolerated. Walk at least 1-2 times in the hall. OOB for more than 8 hours. Up in chair for all meals.<br>For patients on Magnesium Sulfate, mobilization will be limited to sitting on the edge of the bed/ OOB.<br>*After 24h postoperative*<br>Walk at least 3-4 times in the hall. OOB for more than 8 hours. Up in chair for all meals. For patient on magnesium 24-48 hours postoperative: Walk at least 1-2 times in the hall. OOB for more than 8 hours. Up in chair for all meals. After 48 hours postoperative: usual protocol (walk at least 3-4 times in the hall. OOB for more than 8 hours. Up in chair for all meals). | 9 |
| | | (n) Postoperative diet and bowel regimen management | 9 |

**Supplementary Table 1. RECOvER Checklist.**

| | Item | Recommendation | Page |
|---|---|---|---|
| | | Ice chips and/or water within one h of admission to PACU.<br>Chewing gum and advance to a regular diet within four hours post-cesarean as tolerated (modify as appropriate, i.e., diabetic diet for diabetic patients or low sodium diet for patients with renal disease). | |
| | | (o) Criteria for discharge<br>Tolerating PO intake, voiding independently, pain well controlled on oral medication, ambulating in hallways. | N/A |
| | | (p) Tracking of post-discharge outcomes<br>Follow-up appointments were established before discharge. Chart reviews performed by the ERAC team.<br>Obstetrical Transition Care Program contacted patients after discharge. | 18 |
| Enhanced recovery auditing | 12 | ERAC champions were responsible for providing staff support and supervised compliance in the department/ division. With the creation of the ERAC flowsheet, electronic data extraction of the MME use and pain scores and the secondary outcomes were periodically done by the clinical informatics team at the request of the ERAC leaders. This information was used to assess the compliance with documentation and assess the efficacy of the ERAC algorithm. In addition, random charts were reviewed in detail to evaluate compliance with documentation. | 10 |
| Outcomes | 13 | (a) *Primary outcome*<br>Inpatient MME use and the patient's pain scores<br>*Secondary outcomes*<br>Prescribed MME, fasting time for liquids and solids (fasting time before surgery), time to feeding (the time to first feed after arrival to Post Anesthesia Care Unit), time to indwelling urinary catheter removal, time to ambulation, and time to hospital discharge. | 10 |
| | | (b) Clinical outcomes | 11 |
| PROs | 14 | Modified Obstetric Quality of Recovery-10 Scoring Tool. | 7 |
| *Results* | | | |
| Patient population | 15 | See Figure 1 | |
| | | (a) See Table 1 | Table 1 |
| | | (b) Indicate number of participants with missing data for each variable of interest | N/A |

**Supplementary Table 1. RECOvER Checklist.**

| | Item | Recommendation | Page |
|---|---|---|---|
| Enhanced recovery compliance | 16 | Table 2 provides ERAC compliance MME use before and after the implementation. | Table 2 |
| Correlations | 17 | Table 3 provides logistic regression examining MME use with respect to potential confounders. | Table 3 |
| *Discussion* | | | |
| Context | 18 | The study illustrates an ERAC algorithm that reduced inpatient and outpatient opioid use while reducing the pain scores and improving the indicators of postoperative recovery. The algorithm included non-elective CD, used Gabapentin within its multimodal pain regimen. | 15-16 |
| Limitations | 19 | Enrollment of only uncomplicated CD. Performance bias. No verification of opioid prescription dispensed from the pharmacy. No cost-analysis evaluation. | 18 |
| *Other information* | | | |
| Funding | 20 | Support from departmental funds. | N/A |

RECOvER Reporting on ERAS Compliance, Outcomes, and Elements Research; IRB Institutional Review Board; STROBE STrengthening the Reporting of OBservational studies in Epidemiology; PROs patient-reported outcomes.
